# Trans-ethnic analysis of the human leukocyte antigen region for ulcerative colitis reveals shared but also ethnicity-specific disease associations

**DOI:** 10.1101/2020.07.29.20162552

**Authors:** Frauke Degenhardt, Gabriele Mayr, Mareike Wendorff, Gabrielle Boucher, Eva Ellinghaus, David Ellinghaus, Hesham ElAbd, Elisa Rosati, Matthias Hübenthal, Simonas Juzenas, Shifteh Abedian, Homayon Vahedi, Thelma BK, Suk-Kyun Yang, Byong Duk Ye, Jae Hee Cheon, Lisa Wu Datta, Naser Ebrahim Daryani, Pierre Ellul, Motohiro Esaki, Yuta Fuyuno, Dermot PB McGovern, Talin Haritunians, Myhunghee Hong, Garima Juyal, Eun Suk Jung, Michiaki Kubo, Subra Kugathasan, Tobias L. Lenz, Stephen Leslie, Reza Malekzadeh, Vandana Midha, Allan Motyer, Siew C Ng, David T Okou, Soumya Raychaudhuri, John Schembri, Stefan Schreiber, Kyuyoung Song, Ajit Sood, Atsushi Takahashi, Esther A Torres, Junji Umeno, Behrooz Z. Alizadeh, Rinse K Weersma, Sunny H Wong, Keiko Yamazaki, Tom H Karlsen, John D Rioux, Steven R Brant, for the MAAIS Recruitment Center, Andre Franke, for the International IBD Genetics Consortium

## Abstract

Inflammatory bowel disease (IBD) is a chronic inflammatory disease of the gut. Genetic association studies have identified the highly variable human leukocyte antigen (HLA) region as the strongest susceptibility locus for IBD, and specifically DRB1*01:03 as a determining factor for ulcerative colitis (UC). However, for most of the association signal such a delineation could not be made due to tight structures of linkage disequilibrium within the HLA. The aim of this study was therefore to further characterize the HLA signal using a trans-ethnic approach. We performed a comprehensive fine mapping of single HLA alleles in UC in a cohort of 9,272 individuals with African American, East Asian, Puerto Rican, Indian and Iranian descent and 40,691 previously analyzed Caucasians, additionally analyzing whole HLA haplotypes. We computationally characterized the binding of associated HLA alleles to human self-peptides and analysed the physico-chemical properties of the HLA proteins and predicted self-peptidomes. Highlighting alleles of the HLA-DRB1*15 group and their correlated HLA-*DQ-DR* haplotypes, we identified consistent associations across different ethnicities but also identified population-specific signals. We observed that DRB1*01:03 is mostly present in individuals of Western European descent and hardly present in non-Caucasian individuals. We found peptides predicted to bind to risk HLA alleles to be rich in positively charged amino acids such. We conclude that the HLA plays an important role for UC susceptibility across different ethnicities. This research further implicates specific features of peptides that are predicted to bind risk and protective HLA proteins.

## INTRODUCTION

Ulcerative colitis (UC) is a chronic inflammatory disease of the gut. Like Crohn’s disease (CD), the other main subphenotype of inflammatory bowel disease (IBD), it is most likely caused by an abnormal reaction of the immune system to microbial stimuli with environmental factors also playing a role. Currently more than 240 genetic susceptibility loci have been associated with IBD in Caucasians the majority of which are shared between UC and CD (Jostins *et al*., Liu *et al*., Ellinghaus *et al*., de Lange *et al*.^1–4^). Strong genetic association signals with both diseases have been identified in the human leukocyte antigen (HLA) region. The HLA is mapped to the long arm of chromosome 6 between 29 and 34 Mb and moderates complex functions within the immune system. One of the major tasks of the HLA is the presentation of antigens to the host immune system. While HLA class I proteins usually present peptides derived the cytosol (i.e. peptides derived from intracellularly replicating viruses), HLA class II proteins present peptides from extracellular pathogens that have entered the cell e.g. by phagocytosis. In Caucasian IBD patients a large percentage of the phenotypic variation is explained by variants within the HLA class II locus, with DRB1*01:03 being the most significant risk allele for UC (P [P-value]=2.68×10^−119^, OR [odds ratio]=3.59; 95% CI [confidence interval]=3.22-4.00)^5^, specifically by alleles of the HLA-*DR* and -*DQ* loci, though tight structures of linkage disequilibrium (LD) have hindered the assignment of the causal variants. Additionally, a systematic comparison across ethnicities for the HLA association in UC has not been performed, also due to the lack of HLA imputation panels that could accurately infer HLA alleles for trans-ethnic genetic data sets^5–14^. Recently, we created such a trans-ethnic HLA imputation reference panel including dense single nucleotide polymorphism (SNP) fine mapping data typed on Illumina’s ImmunoChip, covering a large proportion of the HLA, within 8 populations of different ethnicities^15^. Here we report the first trans-ethnic fine mapping study of the HLA in UC and some biological implications of the results.

## METHODS

### Cohort description

A detailed description of the cohorts and recruitment sites can be found in the **Supplementary Methods** and **Supplementary Table 1**. In brief, a total of 52,550 individuals (including 18,142 UC patients and 34,408 controls) were used in this study, of which 10,063 (3,517 UC cases and 6,546 controls) were of non-Caucasian origin. The Caucasian, Iranian, Indian and Asian dataset (from which we extracted Japanese and Chinese individuals) are of part of the data freeze published in Liu *et al*.^2^, while individuals of African American (Huang *et al*.^16^), Korean (Ye *et al*.^17^), Maltese, and Puerto Rican descent were added. The recruitment of study subjects was approved by the ethics committees or institutional review boards of all individual participating centers or countries. Written informed consent was obtained from all study participants.

### Genotyping & Quality control

All individuals were typed on the Illumina HumanImmuno BeadChip v.1.0 or the Illumina Infimum ImmunoArray 24 v2.0 (Malta). Genotypes of the study subjects were quality controlled as described in the **Supplementary Methods**.

### Phasing of single nucleotide variants

Using SHAPEIT2^18^ version r727, we phased quality-controlled genotype data on chromosome 6, 25Mb to 34Mb of the respective cohorts using variants with MAF >1%. We excluded SNPs that did not match 1000 Genomes Phase III^19^ (October 2014) alleles (published with the SNP imputation tool IMPUTE2^20,21^) and ATCG variants that did not match the AFR, EUR, SAS, EAS or AMR populations (+ strand assumed for both). AFR (used for comparison with our African American samples), EUR (Caucasian, Iranian, Maltese, SAS (Indian), EAS (Chinese, Korean, Japanese) and AMR (Puerto Rican). Using default values of SHAPEIT2 (--input-thr 0.9, --missing-code 0, --states 100, --window 2, --burn 7, --prune 8, --main 20 and –effective-size18,000, we first generated a haplotype graph and, as suggested by the authors of SHAPEIT2, calculated a value of phasing certainty based on 100 haplotypes generated from the haplotype graph for each population separately. Then, we excluded SNPs with a median phasing certainty<0.8 within each population separately.

### Imputation of single nucleotide variants

To increase the density of single nucleotide variants (SNVs, including variants with minor allele frequency (MAF)<1%) within the HLA region, we used publicly available nucleotide sequences of HLA alleles and further imputed SNVs based on the HLA alleles imputed for each individual using IMPUTE2^22^ with the 1000 Genomes Phase III^19^ individuals as reference (October 2014) using parameters: -Ne 20,000, -buffer 250, -burnin 10, -k 80, -iter 30, -k_hap 500, -outdp 3, -pgs_miss, -os 0 1 2 3, allowing additionally for the imputation of large regions (-allow_large_regions). Imputation of the Caucasian data set was performed in batches of 10,000 samples. Imputation quality control was performed post-imputation excluding variants with an IMPUTE2 info score <0.8. For the Caucasian data set, we excluded variants with a median IMPUTE2 info score <0.8 and a minimum IMPUTE2 info score <0.3. Additionally, we imputed SNPs into the data set using imputed HLA allele information (i.e. translated imputed HLA information into real nucleotide information at each position of the allele) (**Supplementary Methods**).

### HLA Imputation

QC-ed genotype data for each cohort were imputed using Beagle version 4.1^22,23^ based on the corresponding genotype variants observed in the respective cohort. We imputed HLA alleles at loci HLA-*A*, -*C*, -*B, -DRB3, -DRB5, -DRB4, -DRB1, -DQA1, -DQB1, -DPA1* and *-DPB1* at full context 4-digit level using the IKMB reference published in Degenhardt *et al*.^15^ and the imputation tool HIBAG^24^. Imputation of the Caucasian panel was additionally performed with the HLARES panel published with HIBAG (ImmunoChip-European_HLARES-HLA4-hg19.RData). Alleles were not excluded by setting a posterior probability threshold. However, we took the sensitivity and specificity measures we generated as previously reported (Degenhardt *et al*.^15^) into consideration during interpretation.

### Generation of HLA haplotypes

HLA haplotypes were generated by comparing SNP haplotypes generated by SHAPEIT2^18^ for each individual and SNP haplotypes stored for the alleles within the classifiers of the HLA reference model^15^ for the alleles that were imputed for each individual at a given locus. For 10 random classifiers, we calculated the minimal distance between the SNP haplotypes stored for the allele of interest in the HLA reference model and the SNP haplotypes generated by SHAPEIT2. We assigned alleles to a parental haplotype based on how often this allele had minimal difference to the haplotype. Phasing certainty was calculated as the percentage of times an allele was correctly assigned to the chosen parental haplotype. In cases no decision could be made or both alleles were assigned to the same haplotype, phasing certainty was set to 0. If an individual was homozygous at a locus, phasing certainty was set to 1.

### HLA haplotype benchmark

We tested the generation of HLA haplotypes with the above method, using genotype information of trio samples (Utah Residents (CEPH) with Northern and Western European Ancestry (CEU) and Yoruba in Ibadan, Nigeria (YRI)) extracted from the Hapmap Phase 3 project and HLA allele information published for these individuals in the 1000 Genomes HLA diversity panel^25^ using the most common allele for ambiguous calls. In total, 27 CEU samples and 24 YRI samples and their parents were analyzed. Genotype data were downloaded from the HapMap Phase 3 Server (version 2015-05) and positions present on the Illumina ImmunoChip were extracted. We applied the procedure described above for phasing of HLA alleles. The results are shown in **Supplementary Table 2**.

### Calculation of marginal probabilities for each allele

Since HIBAG stores a matrix of all posterior probability values of each (biallelic) allele combination per individual, we calculated the marginal sums of posterior probability for each allele per individual. The overall marginal probability of an allele was then calculated as the mean of the marginal sums of the posterior probability calculated for alleles predicted to carry this allele.

### Association analysis

Subsequently, we performed a standard logistic regression association analysis on single alleles and SNVs. HLA alleles were coded as present (P) or absent (A) with genotype dosages (PP=2, AP=1 and AA=0) by simply counting the number of times an allele occurred for a specific individual. SNPs imputed with IMPUTE2 were included as dosages. SNVs inferred from HLA alleles were coded as 0,1,2 on the minor allele. Additive association analyses for each marker were performed using

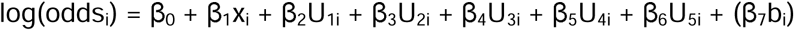

for individual i=1,…..,N, genotype dose or call (x) and eigenvectors (U1-U5). For the analysis of the Puerto Rican and Indian cohort we additionally adjusted for batch (b) (**Supplementary Methods;** batches during QC).

### Meta-analysis

We performed a meta-analysis of association statistics from the 9 analysed cohorts using the tool RE2C^26^. Classical fixed-effects or random-effects meta-analyses are not optimal for the analysis across study estimates where underlying allele frequencies are different between cohorts or similar only for some of the analyzed cohorts (Morris *et al*.^27^) as in the case of trans-ethnic analyses. Using REC2 (Lee *et al*.^26^), a tool optimized for the analysis of heterogenous effects, we combined the association statistics for all 9 cohorts for SNPs and HLA alleles with MAF (SNPs) and AF (HLA alleles) >1% in the respective cohorts. to calculate a combined P-value, setting the correlation between studies to uniform. Here we either report the REC2p* p-value or the REC2 p-value if exceeding heterogeneity was observed for an association. For the original definition see Lee *et al*.^26^.

### Clustering according to preferential peptide binders

Using NetMHCIIpan-3.2^28^, we predicted binding affinities for five sets of the 200,000 unique random 15mer peptides **(Supplementary Methods)** for all alleles that were significant in the meta-analysis and had a frequency of >1% in at least one of the 9 populations. We give the amino acid distribution of these sets in **Supplementary Table 3**. We selected the top 2% (strong binders (SB)) preferential peptide binders as given by the NetMHCIIpan-3.2 software for each allele and calculated the pairwise Pearson correlation between alleles based on complete observations for the respective allele combinations^28^ using R (version 3.3.1), creating a matrix of correlations. Clustering was performed on this matrix using hclust of the R package stats. Correlation between the clusters was calculated using corrplot (version 0.84) and dendextend (version 1.12). Here, the correlation between cluster dendrograms (i.e. the concordance of the tree-structure) is calculated with a value of 0 signifying dissimilar tree-structures and 1 signifying highly similar tree-structures. Dendrograms were plotted using the ape (version 5.3) package, for DQ and DRB1.

### Generation of combined peptide motifs

Based on the clusters generated above for the human peptides, we grouped the risk alleles and protective alleles into 2 clusters each **(Supplementary Methods)**. For each of the 5 peptide sets, we concatenated the top 2% ranked binders (percentile rank of NetMHCIIpan-3.2) for alleles within each protective and risk group and excluded peptides that were among the 10% top ranked binders (percentile rank of NetMHCIIpan-3.2) in two or more of the groups. Based on this, we generated peptide binding motifs using Seq2Logo.^29^ for each of the groups and also plotted the (Position-specific scoring matrix) PSSM scores for chosen amino acids within a group.

### Clustering according to physico-chemical properties

Clustering of HLA proteins was performed using 5 different numerical scores: the Atchley scores F1 and F3^30^, residue-volume^31^ and self-defined parameters charge and hydrogen-acceptor capability **(Supplementary Table 4)**. The amino acid sequence of each respective allele was extracted at positions noted in **Supplementary Table 5** for Pockets 1, 4, 6, 7 and 9 from HLA allele protein sequences that were retrieved from the IMGT/HLA database (version3.37.0)^32^ and aligned using MUSCLE^33^. The alpha chain, of the HLA-*DR* locus is invariable and was not considered in the analysis of this locus. For the respective analysis, each amino acid was assigned its numerical score. Clustering was then performed on the scores using the hclust function of the R (version 3.3.1) package stats and Euclidian distances.

## RESULTS

Here we imputed HLA alleles for a total of 9 cohorts **(Supplementary Figure 1, Supplementary Table 1)** within 3 HLA class I (HLA-*A*, -*C* and -*B*) and 8 class II loci (HLA-*DRB3*, -*DRB5*, -*DRB4*, -*DRB1*, -*DQA1*, -*DQB1*, -*DPA1* and *-DPB1*) at full context 4-digit level utilizing a median of 8,555 SNP genotypes (located within extended HLA between 25 and 34 Mb on chromosome 6p21) from Illumina’s ImmunoChip. After QC, a total of 17,276 UC cases and 32,975 controls remained. 13,927 cases and 26,764 controls were previously reported Caucasians^5^ and 3,251 cases and 6,021 controls were non-Caucasian individuals (Liu *et al*.^2^). After SNP imputation and respective quality control a median of 88,087 SNVs with INFO score > 0.8 were additionally analysed.

In line with our previous study in Caucasians^5^, we observed strong, consistent association signals for SNPs and HLA alleles within the HLA class II region, featuring HLA-*DRB1*, HLA-*DQA1*, and HLA-*DQB1*, for all UC case-control panels except the small-sized Puerto Rican and Maltese cohorts **(Figure 1 and Supplementary Figure 2)**. The strongest association signal was seen for SNP rs28479879 (RE2Cp=5.41×10^−157^, RE2Cp*=8.87×10^−156^, I^2^=79), located in the HLA-*DR* locus, including HLA*-DRB1* and HLA-*DRB3/4/5*. In the Japanese and Korean panels, we further observed a “roof-top”-like association signal spanning the HLA class I and II loci **(Figure 1)** that, as we subsequently demonstrated, was caused by strong LD between the most disease-associated class II alleles DRB1*15:02, DQA1*01:03, and DQB1*06:01, and the class I alleles B*52:01 and C*12:02. The “roof-top”-like signal disappeared when conditioning on class I and class II alleles separately **(Supplementary Figure 3)**. Likely due to lack of statistical power, e.g. for the Maltese data set, and/or diversity of the population, e.g. for the Puerto Ricans, association P-values for these populations did not achieve the genome-wide significance threshold (P<5×10^−8^).

**Figure 1.**
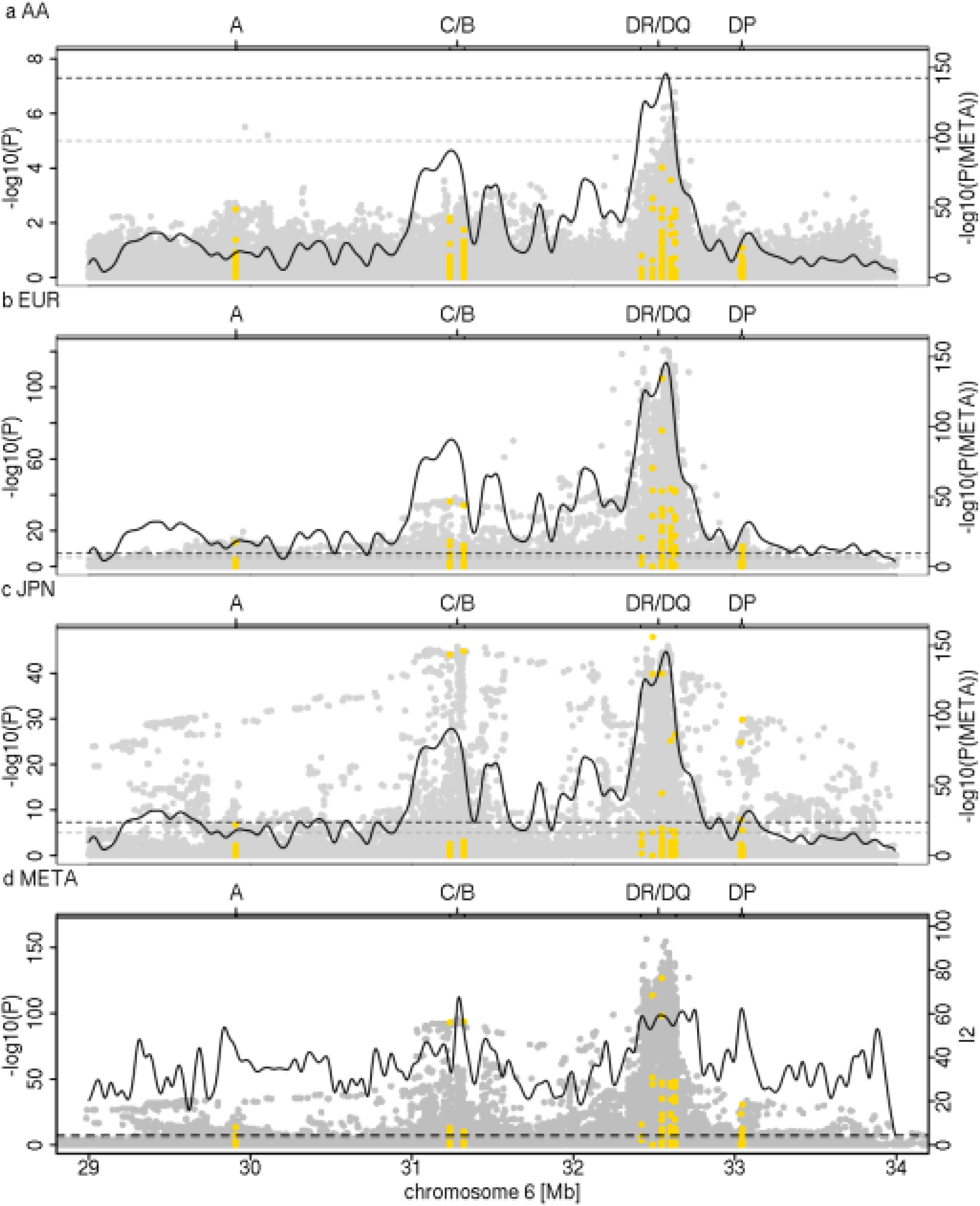
HLA regional association plots. Association analysis results for imputed and genotyped single nucleotide variants (grey) and 4-digit HLA alleles (yellow) are shown for **(a)** 373 African American cases and 590 controls (AA), **(b)** 13,927 Caucasian cases and 26,764 controls (EUR), and **(c)** 709 Japanese cases 3,169 and controls (JPN) as well as **(d)** the meta-analysis (META) results from the analysis with RE2C (Lee *et al*.^26^) at variants with a MAF > 1% in the respective cohorts (including 17,276 cases and 32,975 controls from 9 different cohorts). The association plots for the remaining populations are provided in **Supplementary Figure 2**. SNP. The curves in (a)-(c) show the P-value of the meta-analysis (REC2p* or REC2p). In (d) the overlying curve shows the I^2^ as a measure of heterogeneity in the meta-analysis indicating the heterogeneity of effects and allele frequencies in that region. Dashed lines indicate the thresholds of genome-wide (P=5×10^−8^) and nominal significance (P=10^−5^) The association analyses indicate HLA class II as the most associated susceptibility region across the different populations. In the Korean and the Japanese populations, a strong association signal is also seen for B*52:01 and C*12:02, both alleles being in strong linkage disequilibrium with the HLA class II loci DRB1*15:02, DQA1*01:02 and DQB1*06:01, i.e. another population-specific haplotype association in these ethnicities exists.

The most strongly and consistently associated class II risk alleles within the meta-analysis were alleles of the DRB1*15 group (RE2Cp*=1.87×10^−116^, RE2Cp=1.31×10^−117^, I^2^=92) **(Figure 2, Supplementary Table 6)**, observed to be located on the same haplotype as DQA1*01:02/03 and DQB1*06:01/02 **(Figure 3, Supplementary Table 7)**. DRB1*15:02 was most frequent in the Asian populations (Japanese, Korean), while DRB1*15:03 was specific to the African American population and DRB1*15:01 had the stronger association and higher allele frequency in the Chinese and Caucasian population **(Figure 2, Supplementary Table 6)**, which is consistent to data published in the HLA allele frequency database^34^. Since effect sizes were heterogeneous across populations, we did not compute a combined score, but rather show the OR in **Supplementary Figure 4**. Other associated class II alleles included DQA1*03 alleles (RE2Cp*=5.83×10^−81^) that were observed to be located on a haplotype with DRB1*04 (RE2Cp*=2.36×10^−55^), DRB1*07:01 (REC2p*=5.99×10^−35^, RE2Cp=6.485.99×10^−36^, I^2^=68) or DRB1*09:01 (RE2Cp*=2.73×10^−12^). DRB1*04/07/09 alleles are all located on the same haplotype as HLA-*DRB4* alleles^15^, therefore absence of HLA-*DRB4*, hereafter named DRB4*00:00, was significantly associated with high risk (RE2Cp*=2.35×10^−127^). Along the same line HLA-*DRB5* is located on the same haplotype as DRB1*15. Its absence was therefore observed to be protective. We identified DRB1*10:01 as a novel association signal (RE2Cp*=1.03×10^− 6^). It was observed to be most frequent in the Iranian (3.2% controls and 1.6% cases) and Indian (6.7% controls and 3.3% cases) populations and rare in other populations **(Supplementary Table 6)**, which is most likely why it has not been described before. Among population-specific signals, we also observed significant association of UC with DRB1*14:04 (P=0.004, OR=1.64 95%CI: 1.18-2.29) in the Indian population **(Figure 2)**. Overall, alleles of 11 of the 13 known HLA-*DRB1* 2-digit groups and all 5 known -*DQB1* groups were associated with UC across the different cohorts **(Figure 2, Supplementary Table 6, Supplementary Figure 5)**, with more HLA-*DRB1* alleles conferring protection than risk. Effect sizes in the larger Caucasian and Japanese populations were observed to be moderate (0.5<OR<2.0 for alleles with AF>1%, with the exception of DRB1*15:02 (OR=2.87; 95% CI: 2.46-3.36 in the Japanese population). The comparison of beta estimates also showed that Japanese and Korean effects estimates were most similar (weighted correlation of 0.84, P=1.3×10^−30^), while Iranian and Indian effects estimates correlated better with those of the Caucasian population (weighted correlation of 0.65, P=1.0×10^−16^ and 0.69, P=2.0×10^−17^, **Supplementary Figure 6, Supplementary Methods**). Notably, we identified DRB1*01:03, which was identified as the strongest association signal for IBD in our previous fine mapping analysis^5^ as being population specific. It was not present in the Asian populations and was only observed with a frequency of <0.1% in the African American and Puerto Rican populations. Detailed analysis of the geographic distribution of the DRB1*01:03 allele showed, that it seemingly occurs in Western Europe (Great Britain, Ireland, France, Spain) and former Western colonies with AF >1%, while it seems to be infrequent in the Eastern parts of Europe. We therefore hypothesize that this allele is linked to the history of Western European countries. **(Figure 6)**. Within this study, the frequency of DRB1*01:03 in the Caucasian population is likely underestimated and therefore not the top associated signal in the Caucasian analysis (i.e. DRB1*01:03 was imputed as DRB1*01:01 or DRB1*01:02 due to similarities in SNP haplotype between these alleles) due to applying a reference panel containing mostly non-Caucasian individuals and European individuals from Germany only. Indeed, using the European HLARES imputation panel, which contains a more diverse Caucasian population, we re-established the signal. The frequency of the remaining alleles imputed with our transethnic reference dataset highly correlated with our original study in the Caucasian population **(Supplementary Figure 7)**. Other DRB1*15, for instance DRB1*15:06 did not show association with UC. Interestingly, however DRB1*15:06 has the same amino acid sequence as DRB1*15:01 in the peptide binding groove and may therefore biologically indeed play a role in IBD. With low overall global frequency of the DRB1*15:06 allele, it was not statistically associated with UC. It was most frequent in the Indian population (AF= 2.1%, OR= 1.27, 95%CI [0.77, 2.11]. The theoretical power to detect an effect at the given sample size 1,621, with OR 1.27 and AF 2.1% is estimated to be 0.50 for a significance level of 0.05. This is also true for other alleles listed in **Supplementary Table 8**. The deviation from non-additivity of effects at the HLA locus observed in Goyette *et al*.^5^ could not be replicated in this study (data not shown).

**Figure 2.**
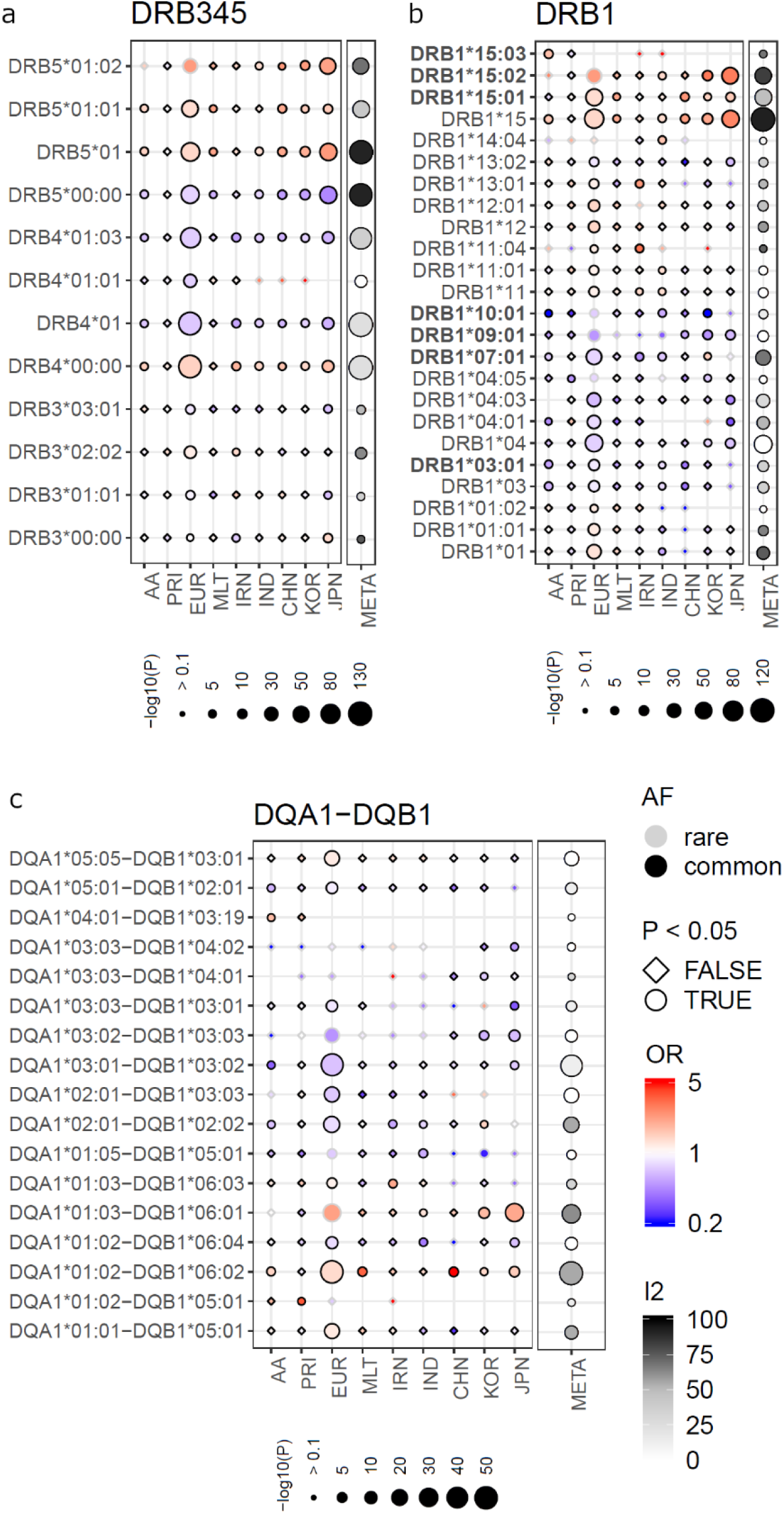
HLA single allele association analysis results at 2- and 4-digit resolution for MHC class II loci *-DRB3/4/5, -DRB1, -DQA1*-*DQB1*. (AF; common defined as AF>1%), odds ratio (OR), P-value (P) and whether an allele had a P-value<0.05 (circle symbol) is shown for the respective population (e.g. circles with black boundary and red color represent an allele that is common and associated with risk). We depict association results of the analysis of the African American (AA), Puerto Rican (PRI), Caucasian (EUR), Maltese (MLT), Iranian (IRN), North Indian (IND), Chinese (CHN), Korean (KOR) and Japanese (JPN) cohorts and the meta-analysis (META) with RE2C’s I2 as an indicator of allelic heterogeneity and the P-value of association (RE2Cp or RE2Cp*, combined here with single study P-values P). Only HLA alleles which are significant in the meta-analysis, that have an AF>1% in at least one population and that have a marginal post imputation probability >0.6 are shown. The strongest association signals in the meta-analysis are for risk alleles of the DRB1*15 group, i.e. DRB1*15:01, DRB*15:02 and DRB1*15:03 and the alleles located on the same respective haplotype **(Figure 3)**. Alleles with OR>5.0 or OR<0.2 (rare and non-significant alleles may have larger/smaller OR) values were “ceiled” at 5.0 and 0.2 respectively. The “consistent alleles” that are highlighted in Figure 3 are highlighted in bold type on the left side.

**Figure 3.**
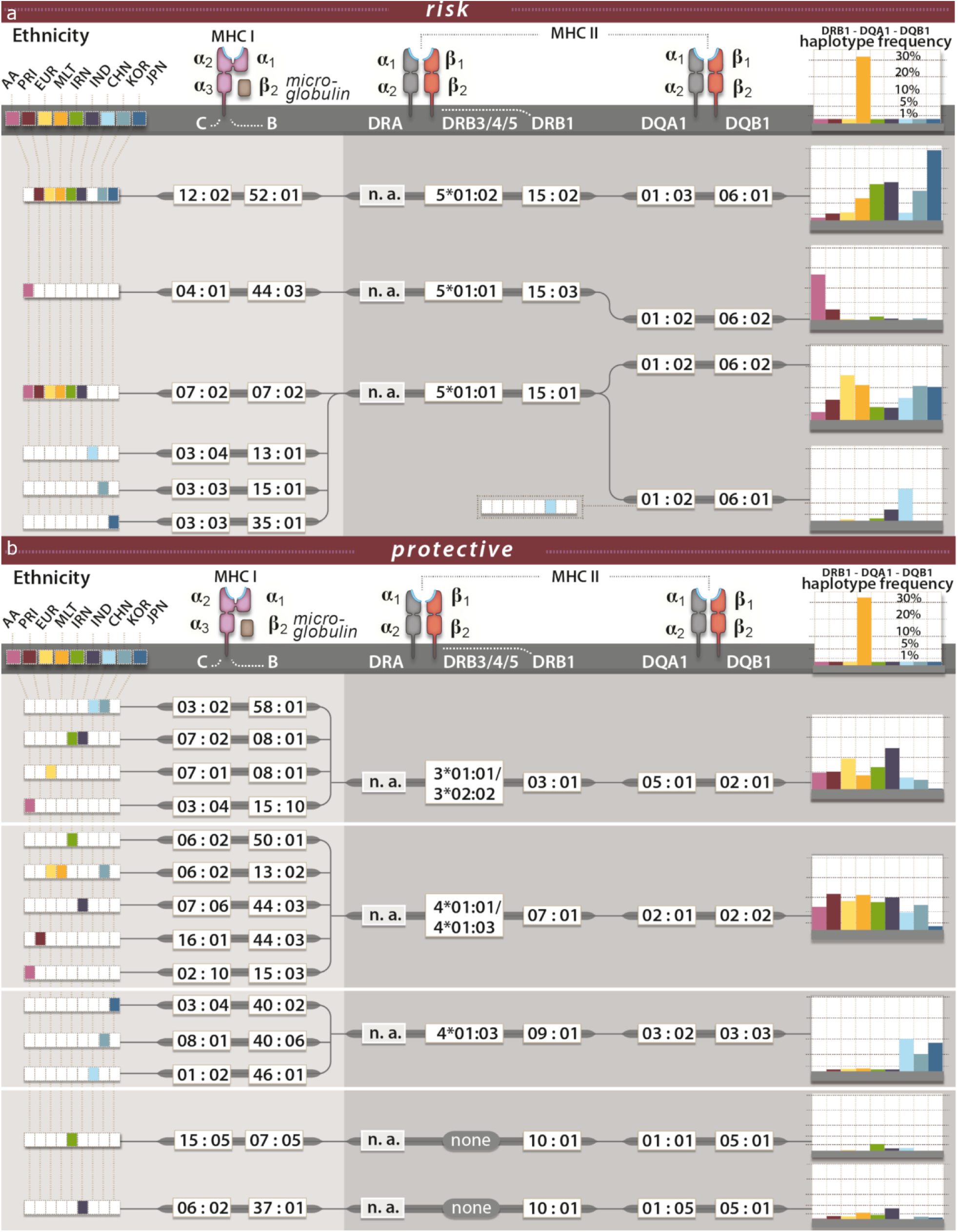
Haplotypes for associated HLA alleles. For a selection of associated HLA alleles, we show the most frequently observed risk **(a)** and protective **(b)** haplotypes in the respective populations. (African American (AA), Puerto Rican (PRI), Caucasian (EUR), Maltese (MLT), Iranian (IRN), North Indian (IND), Chinese (CHN), Korean (KOR) and Japanese (JPN)). Here we show only DRB1-DQA1-DQB1 haplotypes with a frequency >1% in the case individuals in each respective population. The most frequently observed C-B alleles in each population were then added if the C-B-DRB1-DQA1-DQB1 haplotype occurred in more than or equal to 5 individuals. HLA-*DRB3/4/5* alleles were taken from Degenhardt *et al*.^15^ and calculated based on individuals hemizygous for HLA-*DRB3/4/5* (i.e. carrying only one HLA*-DRB1* observed with either HLA-*DRB3*, -*DRB4* or -*DRB5* and one DRB1*01, DRB1*08 or DRB1*10 which are not observed with any of the HLA-*DRB3/4/5*.)

To reduce the complexity of the HLA signal further and to identify the properties of potential culprit antigens leading to disease, we analysed peptides preferentially bound by proteins, attributed risk and protection on the genetic level **(Figure 4)**. Additionally, we tried to identify shared physico-chemical properties of these proteins. For this analysis, we only selected proteins for which the corresponding alleles had a significant P-value in the meta-analysis (RE2Cp*<0.05) and focused on the results of the DRB1 proteins **(DQ shown in Supplementary Figure 8)**. First, we predicted the binding affinities for 5 sets of 200,000 random unique peptides sampled from the human proteome to the DRB1 proteins using NetMHCIIpan-3.2^28^ (**Supplementary Table 3**, amino acid distribution). Next, we performed clustering analysis on the alleles using the top 2% ranked preferentially binding peptides for each allele based on pairwise observed complete observations. In general, we found DRB1-clustering **(Figure 4)** to be more informative regarding separation of protective and risk alleles than DQ-clustering. Additionally, DRB1-clustering was more stable across the sets of random peptides **(Supplementary Figure 8)**. Larger “risk clusters” were identified for DRB1 including DRB1*15:01 and the newly identified DRB1*15:03. We defined 2 risk clusters including DRB1*11:01/04 and DRB1*13:01 (RISK 1) DRB1*12:01, DRB1*14:04 and DRB1*15:01/03 (RISK 2), and 2 protective clusters including DRB1*04:01/05, DRB1*07:01, DRB1*09:01 and DRB1*10:01 (PROT 1) and DRB1*04:03/06 (PROT 2). Within each cluster, we calculated a unique peptide binding motif by combining the top 2% of binders for each allele in the groups **(Figure 4)**. The peptide binding motifs of the two risk groups were enriched for basic amino acids (K and R) and depleted for acidic amino acids, while the peptide binding motifs of the protective group were enriched for hydrophobic and polar amino acids. Interestingly, DRB1*01:03 clustered with protective alleles DRB1*04:01/05, DRB1*07:01, DRB1*09:01 and DRB1*10:01, however, a more detailed analysis of its physico-chemical properties resulted in a predominant clustering with DRB1*15 **(Figure 5)**. Equally, DRB1*15:02 clustered with DRB1*13:02, while physico-chemical properties resulted in a predominant clustering with the DRB1*15 group. In **Supplementary Figures 9-12** we show that this may be an artefact of NetMHCIIpan-3.2 caused by extrapolation of the DRB1*15:02 signal for unknown peptides from DRB1*13:02.

**Figure 4.**
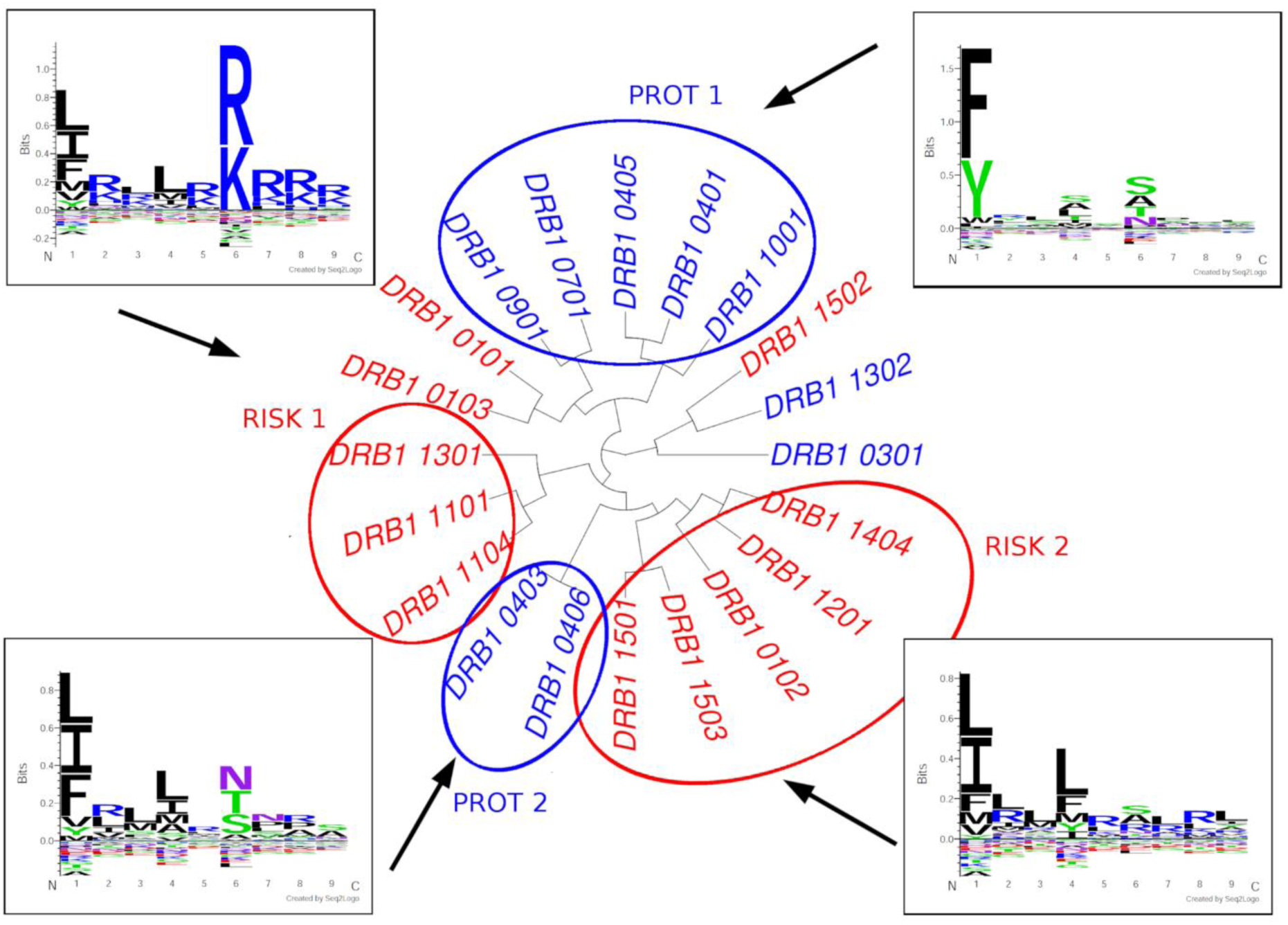
Clustering of DRB1 proteins according to preferential peptide binding and combined peptide binding motifs. (MIDDLE CLUSTER): For 5 sets of 200,000 unique random human peptides the percentile rank scores of preferential peptide binding were calculated using NetMHCIIpan-3.2.^28^ for all DRB1 proteins that were significant in the meta-analysis of genetic analysis of the HLA with and AF > 1% in at least one cohort. We additionally included DRB1*01:03. Within each set, the top 2% binders (according to NetMHCIIpan-3.2 threshold) were used to perform a clustering on the pairwise correlations between two alleles using complete observations only. We show clustering results for peptide set 2. Labels were colored according to risk (red) or protective (blue). (BINDING MOTIFS): Top 2% binders were combined for proteins (RISK 1) DRB1*11:01/04 and DRB1*13:01 DRB1*12:01, DRB1*14:04 and DRB1*15:01/03 (RISK 2), DRB1*04:01/05, DRB1*07:01, DRB1*09:01 and DRB1*10:01 (PROT 1) and DRB1*04:03/04/06 (PROT 2). For this analysis shared peptides (10% top binders) between at least two of the groups where deleted from the set. Here we depict the results for human peptide set 2. Peptide motifs were plotted using Seq2Logo.^29^. The color scheme shows the chemistry of the amino acids. *Red*: positively charged amino acids, *blue*: negatively charged amino acids, *green*: polar amino acid, *purple*: neutral amino acid and *black*: hydrophobic amino acid.

**Figure 5.**
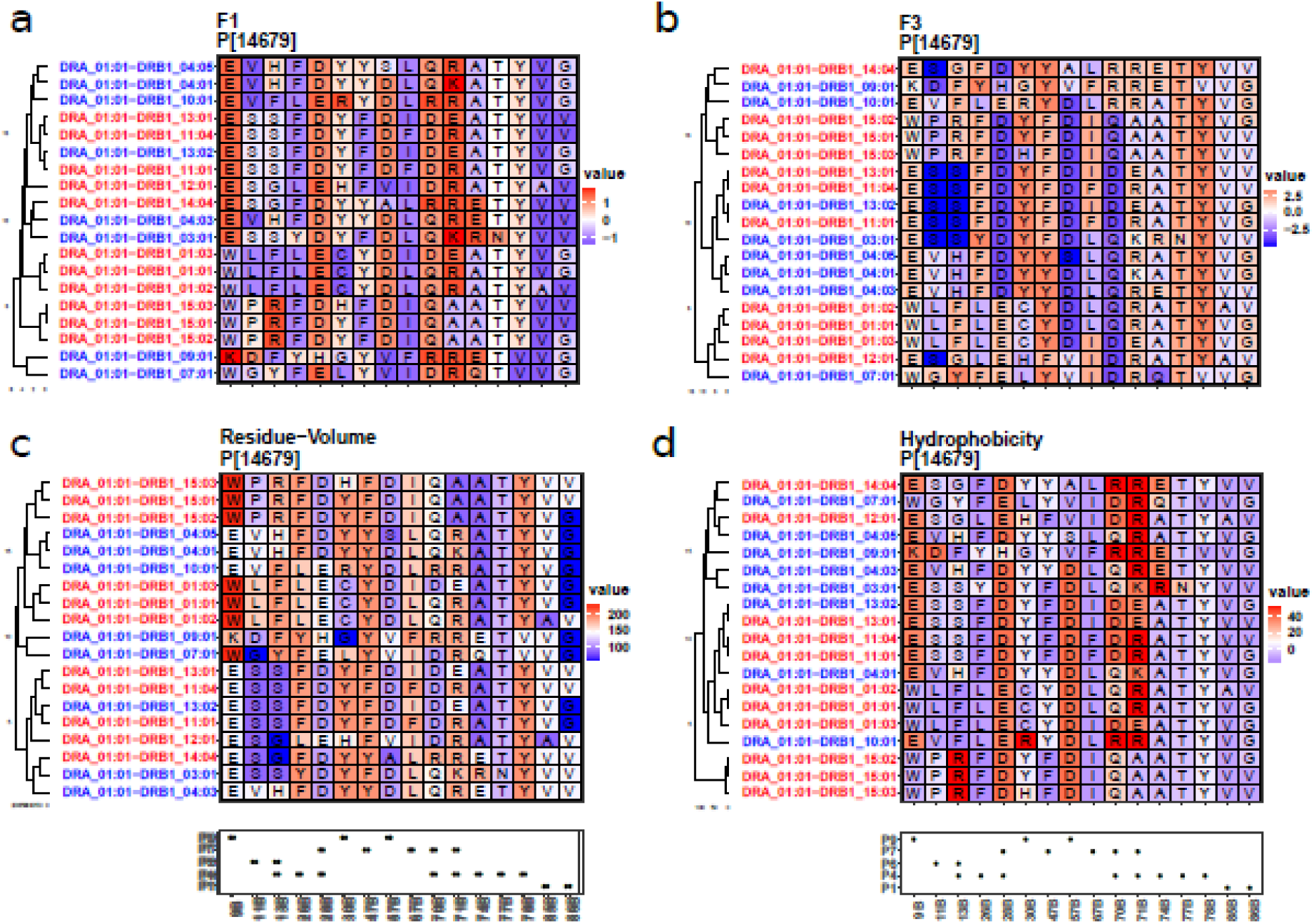
Cluster according to chosen physico-chemical properties of amino acids within the peptide binding pockets. We only show sites with variable information in pockets (P) 1, 4, 6, 7 and 9 and only proteins for which the genetic analysis was significant (meta-analysis RE2Cp/RE2Cp* <0.05) and for which at least 1 cohort had AF >1%. We additionally show DRB1*01:03. Clustering was performed using the hclust function of the R package stats. The box below the cluster plot shows positions of P1, 4, 6, 7 and 9 of the beta (B) chain of the molecules (as defined in **Supplementary Table 5**). Here we show combined scores F1 **(a)** and F3 **(b)** derived from a factor analysis of 54 unique amino acid properties (Atchley *et al*.^30^). F1 captures polarity and hydrophobicity of the amino acid, while factor F3 captures amino acid size and bulkiness. For F1, high values indicate larger hydrophobicity, polarity and hydrogen donor abilities while low values indicate non-polar amino acids. For F3, high values indicate larger and bulkier amino acids while low values indicate smaller, more flexible amino acids. We additionally show the residue-volume **(c)** as a measure of pocket size and defined a score “hydrogen acceptor” (HB-acceptor) **(d)**, which defines the ability of an amino acid to participate in hydrogen bonds and corresponds to the number of atoms within the sidechain that can accept a hydrogen. Additional information for the “charge” parameter and the analysis for DQA1-DQB1 can be found in **Supplementary Figures 9**,**10**.

**Figure 6.**
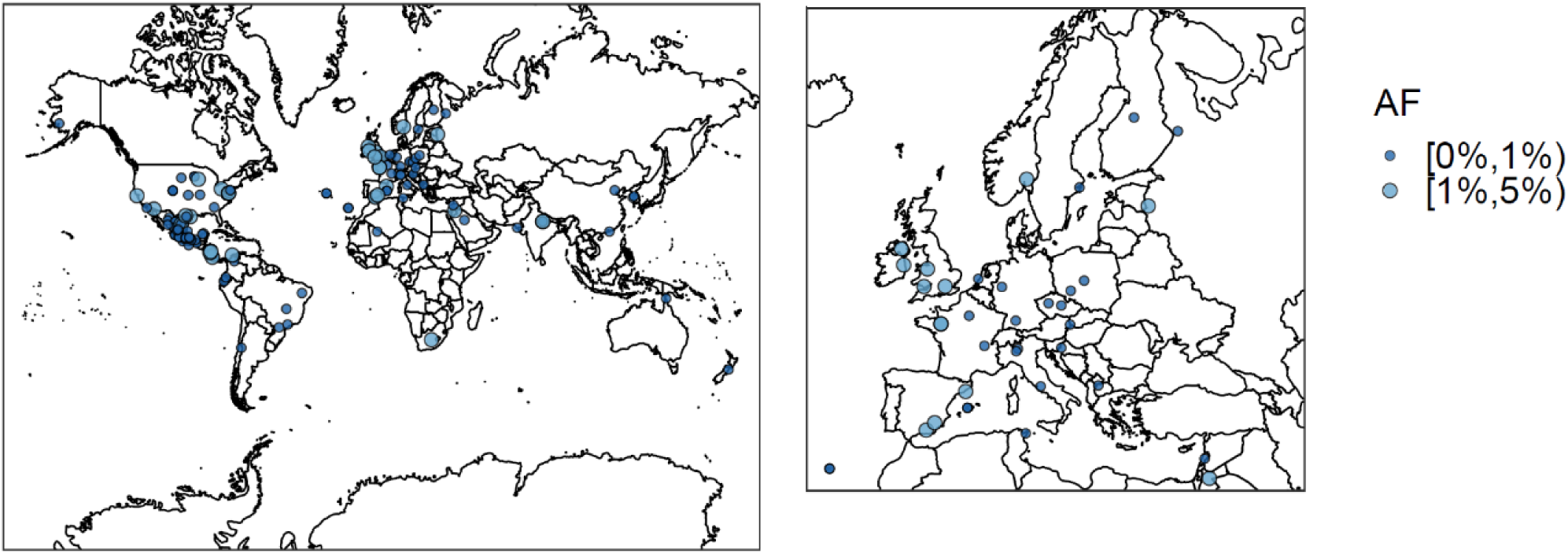
Frequency of DRB1*01:03 across populations available in the allele frequency net database. **(a)** “Worldmap”, **(b)** zoom into European continent. Frequencies are shown within different ranges noted by AF. Allele frequencies of DRB1*01:03 are lower across central Europe than in the UK, Spain, India, South Africa, United States, and coastal regions of South America. Frequencies were binned according to allele frequency. The figures were created using the R-package rworldmap. Frequencies were extracted from the allele frequency network database^34^ for populations larger than 100 individuals. To plot the geographic locations, we converted assigned degree and minutes to decimal numbers. We deleted all non-Caucasian populations with USA coordinates prior to plotting.

## DISCUSSION

Several conclusions can be drawn from this trans-ethnic HLA fine mapping study in UC: HLA allele associations and their effect directions are broadly consistent across the different populations analysed in this study and signals previously observed in a Caucasian-only approach can be replicated in this context^5^. While not in every case the same HLA allele is implicated across the different populations, alleles of the same HLA allele group are associated with UC, as is the case for the HLA allele group DRB1*15, of which DRB1*15:01, DRB1*15:02 and DRB1*15:03 are all associated with the disease dependent on the HLA allele frequencies in each respective population. The frequencies for these alleles computed in this study were consistent to the frequencies stored in the allele frequency net database (AFND) ^34^. Heterogeneity of effect sizes was observed, however the accuracy of estimation of the effect sizes would increase with larger per-population sample sizes. As observed also in the Caucasian-only approach, HLA associations are correlated across different HLA genomic loci, especially for HLA-*DRB1*, -*DRB3/4/5* and -*DQ* alleles, such that neither locus can be ruled out as disease-relevant. Overall a high conservation of HLA-DQ-DR haplotypes was observed across different ethnicities. In the Japanese and Korean population and entire haplotype spanning class I and class II was observed for C*12:01-B*52:02-DRB5*01:02-DRB1*15:02-DQA1*01:03-DQB1*06:01. For South Western Asian (Iranian, Indian) individuals, other HLA associations were observed to be more dominant (i.e. HLA-DRB1*11 and HLA-DRB1*14). Overall, for HLA-association was dependent on the frequency of the allele and the size of the study cohort (i.e. alleles with a sufficiently high frequency at the *DRB1* locus were usually also associated with the disease, except for alleles of the HLA-DRB1*08 and HLA-DRB1*16 groups which had frequency of 2.9% and 1.7% respectively in the Caucasian population). Associations at DPA1-DPB1 can most likely be ruled out and associations may merely result from correlation with HLA-DQ-DRB1. In the analysis of peptide-binding preferences for HLA-*DRB1* alleles, we observed clustering according to the effect’s direction in the genetic analysis, i.e. protective or risk, which may point more to HLA-*DRB1* playing a role. However, an important limitation for the analogous DQ analysis is the limited availability of data present in models for HLA-peptide binding prediction. The highly similar binding pockets of HLA-DRB1*13:01 and HLA-DRB1*13:02 suggest HLA-DQ alleles to mediate disease risk. DRB1*13:01, which was estimated to confer risk is correlated with DQA1*01:03-DQB1*06:03 while DRB1*13:02, which was estimated to be protective, is correlated with DQA1*01:02-DQB1*06:04.

Alleles of the DRB1*15 group also play a role as risk factors in other immune-related diseases including Multiple Sclerosis^35–38^ (a chronic inflammatory neurological disorder), Systemic Lupus Erythematosus^39^ and Dupuytrien’s disease^40,41^ (both are disorders of the connective tissue). They have also been reported to be associated with adult onset Still’s disease^42^ (a systemic inflammatory disease), Graves disease (an autoimmune disease that affects the thyroid), pulmonary tuberculosis and leprosy^43,44^ (a disease caused by *Mycobacterium leprae* that affects the skin). For Multiple Sclerosis, DRB1*15:03, like in our study, was observed to be specific for African American populations^35^. DRB1*15 alleles have been reported to be strongly protective in type 1 diabetes (T1D) (in which the autoimmune system attacks insulin producing beta cells of the pancreas), and pemphigus vulgaris (a skin blistering disease). However, the functional consequences of HLA-DRB1*15 association with these diseases have not been addressed and for most of them the potential disease driving antigens are not known. Exceptions are leprosy, in which *Mycobacterium leprae* causes the disease and pemphigus vulgaris, in which the skin protein desmoglein is targeted. Krause-Kyora *et al*. found that DRB1*15:01, among 18 contemporary DRB1 proteins, was predicted to “bind the second smallest number of potential *Mycobacterium leprae* antigens” and further hypothesized that limited presentation of the *Mycobacterium leprae* antigens, may impair the immune response against this pathogen^43^. Here an important note should be, that DRB1*15:01 is on average also the most frequent HLA-*DRB1* allele in the most analysed British/Central American European populations as such has a higher statistical power to be detected in an association analysis.

Analysis of peptide binding motifs showed that protective and risk alleles cluster stably and that risk and protective groups have peptide binding motifs which are distinguishable by their physico-chemical properties. The arginine (R) and lysine (K) content was observed to be increased in peptides bound by HLA-proteins that were assigned to confer risk on a genetic level. This was more prominent for risk cluster 1 than risk cluster 2. Interestingly, Dhanda *et al*., who compared 1,032 known T-cell epitopes from 14 different sources (including Mycobacterium tuberculosis, Dengue Fever, Virus, Zika Virus, house mite and other allergens) and known non-epitopes from the same data set, showed that T-cell epitope amino acid motifs also are enriched in lysine and arginine content. The established motif is especially similar to the binding motif of risk cluster 1. Arginine is also found at an increased level in antimicrobial peptides^45–47^. Antimicrobial peptides are made of cationic residues and are part of the innate immunity. They target the cell wall of bacteria or structures in the cytosol of bacteria^48^. If and how this plays a role in the etiology of UC is however only to be speculated about.

One important limitation of the analysis of preferential HLA-peptide binding is the amount of data that is used to train machine learning algorithms, which was especially limited for the HLA-DQ proteins. In the future, larger datasets from peptidomics experiment will likely increase the accuracy of these predictions and increase confidence in the risk and protective motifs that may be indicative of culprit antigens in UC due to distinct features. Larger per-population patient collections will be needed in future studies to confirm our results and to obtain even more precise effect estimates of associated HLA alleles. In addition, we hope that IBD patient panels from other ethnicities will become available for genetic fine mapping studies. With typing of HLA alleles now being possible using next-generation sequencing methods, real typing rather than imputation analyzes should become standard, thereby avoiding possible imputation artefacts. The construction of haplotype maps will then likely be even more accurate.

### Description of Supplemental Data

Supplementary Data contain **9 Tables** and **12 Figures**.

## Data Availability

The ImmunoChip data used in this study are proprietary to the IIBGDC genetics consortium and may be requested from the consortium. Any data produced within this study, may be requested from the corresponding authors upon reasonable request including association statistics of imputed and genotyped SNVs.

## Acknowledgements & Grant support

This project received infrastructure support from the DFG Excellence Cluster No. 306 “Inflammation at Interfaces”. M.W. and H.E. are supported by the German Research Foundation (DFG) through the Research Training Group 1743, “Genes, Environment and Inflammation”. E.E. received funding from the European Union Seventh Framework Program (FP7-PEOPLE-2013-COFUND; grant agreement No. 609020 (Scientia Fellows)). S.A. is supported by joint funding from the University Medical Center Groningen, Groningen, The Netherlands, and Institute for Digestive System Disease, Tehran University of Medical Sciences, Tehran, Iran. Funding for the Multicenter African American IBD Study (MAAIS) samples, for the GENESIS samples, and for the African Americans recruited by Cedars Sinai was provided by the U.S.A. National Institutes of Health (NIH) grants DK062431 (S.R.B.), DK 087694 (S.K.), and DK062413 (D.P.B.M), respectively. This work was supported by a grant from the BioBank Japan Project and, in part, by a Grant-in-Aid for Scientific Research (B) (26293180) funded by the Ministry of Education, Culture, Sports, Science, and Technology, Japan. This research was supported by a Mid-career Researcher Program grant through the National Research Foundation of Korea to K.S. (2017R1A2A1A05001119), funded by the Ministry of Science, Information & Communication Technology and Future Planning, and a grant of the Korea Health Technology R&D Project through the Korea Health Industry Development Institute (KHIDI), funded by the Ministry of Health & Welfare (grant number: HI18C0094), Republic of Korea. Funding for the Indian samples was provided by the Centre of Excellence in Genome Sciences and Predictive Medicine (Grant # BT/01/COE/07/UDSC/2008) from the Department of Biotechnology, Government of India). The funders had no role in study design, data collection and analysis, decision to publish, or preparation of the manuscript.

## Author information

Steve R Brant, Tom H Karlsen, John D Rioux and Andre Franke: These authors jointly supervised this work.

## International IBD Genetics Consortium

A full list of members and affiliations appears in the **Supplementary Note**.

## MAAIS Recruitment center

A full list of members and affiliations appears in the **Supplementary Note**.

## Authors contributions

F.D. performed statistical and computational analysis, G.B. contributed to statistical analysis. M.W. and H.E. performed computational analysis with contributions from D.E., M.Hü, S.L., A.M., T.L. and S.R.. G.M. performed protein structure analysis and analysis of physico-chemical properties with contributions from F.D. F.D. and M.W. set up the HLA imputation pipeline. S.J. performed HLA typing in contribution to the HLA reference panel. F.D., G.M., E.E., E.R. wrote or revised this manuscript. S.A., B.A., T.B.K., S-K.Y., B.D.Y., J.H.C., L.W.D., N.E.D., P.E., M.E., Y.F., D.P.B.M., T.H., M.Ho., G.J., E.S.J., M.K., S.K., R.M., V.M., S.C.N., D.T.O, J.S., S.S., K.S., A.S., A.T., E.A.T, J.U., H.V., R.K:W.,S.H:W., K.Y. were involved in study subject recruitment, contributed genotype data and or/phenotype data. F.D., T.H.K., J.D.R., S.R.B. and A.F. conceived, designed and managed the study. All authors reviewed, edited and approved the final manuscript.

## Code availability

Code used for analysis of data within this study is available from upon reasonable request. Part of the code has been incorporated into a github project and is available at ikmb/HLApipe. This pipeline was developed by Frauke Degenhardt and Mareike Wendorff. It is based on the HLA imputation tool HIBAG published by Zheng *et al*.^24^.

## ABBREVIATIONS

(A): Alpha chain of an HLA protein
(B): Beta chain of an HLA protein
AA: African American population of this study
AFR: African American population of the 1000 Genomes/HapMap population see also https://www.internationalgenome.org/category/population/)
AMR: Admixed American population of the 1000 Genomes/HapMap population (see also https://www.internationalgenome.org/category/population/)
AF: Allele Frequency
CEU: Utah Residents (CEPH) with Northern and Western European Ancestry of the 1000 Genomes/HapMap population (see also https://www.internationalgenome.org/category/population/)
CI: Confidence Interval
EAS: East Asian population of the 1000 Genomes/HapMap population (see also https://www.internationalgenome.org/category/population/)
EUR: Caucasian population of this population or (mentioned within the context of the 1000Genomes/HapMap population European data of the latter; see also https://www.internationalgenome.org/category/population/)
F1, F3: Atchley Factors 1 and 3, that contain information on 54 amino acid properties
HLA: Human Leukocyte Antigen
HLA-*A*: Human Leukocyte Antigen gene locus *A*
HLA-*B*: Human Leukocyte Antigen gene locus *B*
HLA-*C*: Human Leukocyte Antigen gene locus *C*
HLA-*DRA*: Human Leukocyte Antigen gene locus *DRA*
HLA-*DRB1*: Human Leukocyte Antigen gene locus *DRB1*
HLA-*DRB3*: Human Leukocyte Antigen gene locus *DRB3*
HLA-*DRB4*: Human Leukocyte Antigen gene locus *DRB4*
HLA-*DRB5*: Human Leukocyte Antigen gene locus *DRB5*
HLA-*DQA1*: Human Leukocyte Antigen gene locus *DQA1*
HLA-*DQB1*: Human Leukocyte Antigen gene locus *DQB1*
HLA-*DPA1*: Human Leukocyte Antigen gene locus *DPA1*
HLA-*DPB1*: Human Leukozyten Antigen gene locus *DPB1*
IND: Indian population
IRN: Iranian population
JPN: Japanese population
KOR: Korean population
MAF: Minor Allele Frequency
MLE: Maximum Likelihood Estimator
MLT: Maltese population
PRI: Puerto Rican population
P1-P9: Pockets 1 to 9 of the HLA protein within the HLA peptide binding site
QC: Quality Control
SAS: South Asian population of the 1000 Genomes/HapMap population (see also https://www.internationalgenome.org/category/population/)
SNP: Single Nucleotide Polymorphism (MAF >= 1%)
SNV: Single Nucleotide Variation (MAF < 1%)
xHLA: extended HLA region
YRI: Yoruba in Ibadan, Nigeria population of the 1000 Genomes/HapMap population (see also https://www.internationalgenome.org/category/population/)

